# Statistical considerations for drawing conclusions about recovery

**DOI:** 10.1101/19013060

**Authors:** Keith R. Lohse, Rachel L. Hawe, Sean P. Dukelow, Stephen H. Scott

## Abstract

**Background:** Numerous studies have found associations when change scores are regressed onto initial impairments in people with stroke (slopes ≈ 0.7). However, there are important statistical considerations that limit the conclusions we can draw about recovery from these studies.

**Objective:** To provide an accessible “check-list” of conceptual and analytical issues on longitudinal measures of stroke recovery. Proportional recovery is an illustrative example, but these considerations apply broadly to studies of change over time.

**Methods:** Using a pooled dataset of N = 373 Fugl-Meyer Assessment (FMA) upper extremity scores, we ran simulations to illustrate three considerations: (1) how change scores can be problematic in this context; (2) how “nil” and non-zero null-hypothesis significance tests can be used; and (3) how scale boundaries can create the illusion of proportionality, while other analytical procedures (e.g., post-hoc classifications) can augment this problem.

**Results:** Our simulations highlight several limitations of common methods for analyzing recovery over time. Critically, we find that uniform recovery (in the population) leads to similar group-level statistics (regression slopes) and individual-level classifications (into fitters and non-fitters) that have been claimed as evidence for the proportional recovery rule.

**Conclusions:** Our results highlight that one cannot identify whether proportional recovery is true or not based on commonly used methods. We illustrate how these techniques (regressing change scores onto baseline values), measurement tools (bounded scales), and post-hoc classifications (e.g., “non-fitters”) can create spurious results. Going forward the field needs to carefully consider the influence of these factors on how we measure, analyze, and conceptualize recovery.

## Introduction

Recently, much ink has been spilt on the topic of the proportional recovery rule in stroke rehabilitation^1^. In its broadest sense, the proportional recovery rule posits that the amount of recovery patients are likely to have is roughly 70% of the total possible recovery they could make, on average, after the exclusion of “non-fitters” to the rule^2,3^. This relationship is usually demonstrated by regressing change scores (a terminal assessment minus the baseline assessment) onto the initial amount of impairment. Not surprisingly, severely impaired individuals show the greatest variation in their potential for recovery, and severely impaired individuals who do not recover very much are classified as “non-fitters” to the general rule.^1,4^ Classification of non-fitters has been based on different methods^4^ that rely on either a statistical classification (e.g., outlier detection^1^), or physiologically relevant outside variables (e.g., cortico-spinal tract integrity^5^) and behavioral tests (e.g., specific items related to distal upper extremity function^6^).

However, there are important statistical considerations we need to make when recovery is quantified in this way (i.e., the calculation and use of change scores, the interpretation of the null-hypothesis significance test, and the validity of the non-fitter classification). Past critiques of proportional recovery have focused especially on the problems with regressing change scores onto baseline impairment and concerns with the sub-group analysis of fitters and non-fitters in some statistical detail^7,8^. A very short summary of these critiques is that data showing proportional recovery are influenced by statistical artifacts and, at the very least, overstated.

In response, Kundert et al^4^ authored a rebuttal in favor of the proportional recovery rule. Kundert and colleagues’ response incorporates some previous critiques and seeks to refute other criticisms in their discussion, ultimately concluding that proportional recovery is a real biological phenomenon and representative of spontaneous recovery. In their abstract, Kundert and colleagues conclude that, “existing data in aggregate are largely consistent with the [Proportional Recovery Rule] as a population level model for upper limb motor recovery; recent reports of its demise are exaggerated, as these excessively focus on the less conclusive issue of individual subject level predictions.” Those authors also write, “new analytical approaches will be needed to confirm (or refute) a systematic character to spontaneous recovery […] which can be captured by a mathematical rule either at the population or at the subject level.” In this point of view, we argue that Kundert et al.’s^4^ first assertion is not correct, but we echo their second statement that new analytical approaches are needed to confirm (or refute) the systematic character of recovery following stroke.

Below, we critique the evidence in favor of the proportional recovery rule based on three statistical considerations. Using simulations, we illustrate these problems visually. We hope that this simulation-based approach makes the critique more intuitive and accessible to a general audience. Note that these considerations apply to recovery at the “population level”, but we will also discuss the issue of individual prediction and how individual/aggregate data relate. Our three statistical consideration are:

1. **The calculation of change scores is problematic, especially when regressed onto baseline values**. Simple difference scores have long been regarded as a sub-optimal method for assessing change over time^9,10^. Although there are cases where change scores are valid, they are generally inferior to statistically “controlling for” baseline assessments as a covariate. In the case of proportional recovery, an additional hazard is created because change scores are being regressed onto baseline scores, which creates a mathematical coupling^11-13^.
2. **It is important to reflect on what the null-hypothesis test of a regression slope really means, to consider appropriate null hypotheses, and what alternative explanations remain**. To say that a regression slope is statistically significant (e.g., *b* ≈ 0.7, *p* < 0.05), means that if we assume the null hypothesis is true and all of our assumptions hold, we would expect to get a slope greater than or equal to the observed slope less than 5% of the time. However, we need to consider the appropriate null hypotheses against which to test (e.g., we commonly assume a true effect = 0, but we could select other values) and, if we reject the null, we need to consider which alternative explanations remain on the table.
3. **Scale boundaries can create the illusion of proportionality (e.g., floor/ceiling effects) and other analytic steps may augment this problem (e.g., spurious identification of “non-fitters”)**. Using simulations informed by empirical data, we can show what we would observe if the underlying change is random under a uniform distribution, rather than proportional. Using hierarchical cluster analysis to identify non-fitters in our simulations, we can show that data from N=373 real stroke patients on the Fugl-Meyer Assessment is consistent with random uniform recovery. As such, current data do not support the claim that recovery is proportional any more than that recovery is uniform.

We stress that proportional recovery is the motivating example here, but these considerations apply to the study of recovery broadly. Recovery is a difficult problem and choices made in design, measurement, and statistical analysis can either make that problem clearer or can obfuscate the issue.

In the Discussion, we focus on some of the positive evidence from the proportional recovery literature and suggest productive ways to move forward analytically. For instance, neuroanatomical differences do have strong associations with the potential for recovery at different levels of impairment^5,14,15^. However, regressing change scores onto baseline scores is rife with statistical problems. We recommend that if researchers want to explain individual differences in recovery over time, then we should be using formal conditional longitudinal models with more data points and avoiding the statistical confounds of change scores.^16,17^ Indeed, a number of researchers have started making strides in this direction, using longitudinal methods to explore trajectories of stroke recovery^18,19^. Understanding which factors explain, or better yet predict^20-22^ stroke trajectories is a very important area of research.

## Consideration 1: The use of change scores is problematic

Difference scores have been critiqued for many years in the biomedical literature as a method for capturing change^9,10,23^. The reason for this is that difference scores implicitly assume a one-to-one relationship between pre-test scores and post-test scores. This implicit assumption can be seen more clearly if we contrast the formula for a linear regression controlling for baseline (Eq 1.) against a linear regression in which difference scores are the outcome (Eq 2.):

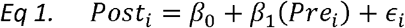

In Eq 1, the relationship between pre-test scores and post-test scores is weighted based on the correlation between time-points in the data (ultimately creating the regression coefficient *β*_1_).

In contrast, if difference scores were our outcome, we would have a formula like Eq 2A:

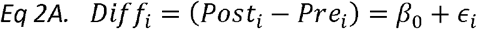

To see the correspondence between controlling for pre-test as a covariate (Eq 1) and treating difference scores as an outcome (Eq 2A), we can simply move our pretest scores to the other side of the equals sign (Eq 2B):

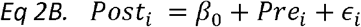

Note that there is not a slope coefficient next to the pre-test variable in Eq 2B, but the implied slope is 1. This slope is implied because in this equation every 1-unit change in pre-test scores, we will have a 1-unit change in post-test scores. (Equation 2B could be equivalently written as 1 *(*Pre_i_*).) Thus, using difference scores as our outcome is equivalent to assuming that the relationship between pre-test and post-test scores is: *β*_1_=1.

Assuming a one-to-one relationship between pre-test and post-test scores might be reasonable when the correlation between pre-test and post-test score is very high, but in general it is much better practice to control for pre-test as a covariate^23^. Controlling for pre-test allows for regression to the mean whereas difference scores do not. That is, random error for lower scoring participants is likely to drive their scores upward on a second measurement, and vice versa for high scoring participants. Regression to the mean is less of a concern if we are dealing with clinical tests with high reliability (because measurement errors from test to test should be small). Even in that case, however, another benefit is that controlling for pretest allows for to be weighted based on the correlation between pre-test and post-test, whereas taking a difference score does not. This is important because when the correlation between pre-test and post-test is low, taking difference scores can actually add noise to the data^9^.

Using change scores is already suboptimal, but the additional step of regressing change scores onto baseline measures leads to the issue of mathematical coupling discussed by Hawe et al.^7^ and Hope et al. ^8^. We illustrate the negative effects of mathematical coupling in Figure 1, using both a normal distribution (1A/B) and a uniform distribution with clear boundaries (1C/D). (See Supplemental Appendix I for all simulation and analysis code.) The point of this illustration is to show that mathematical coupling is a different effect from “boundaries” on a scale, although the two can be related when floor/ceiling effects are present. In both simulations (N = 1000 data points), the variables *X* and *Y* are totally independent (*r*=0.0). However, when we calculate a new variable *Z* =*Y - X*, we find that *Z* and *X* have a strong negative relationship (*r*=-0.7). The reason for this is that *Z* and *X* are mathematically “coupled”; that is, *Z* contains *X* so they are intrinsically linked. This can be seen a little more clearly if we rearrange the terms for in a regression equation:

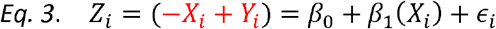

**Figure 1.**
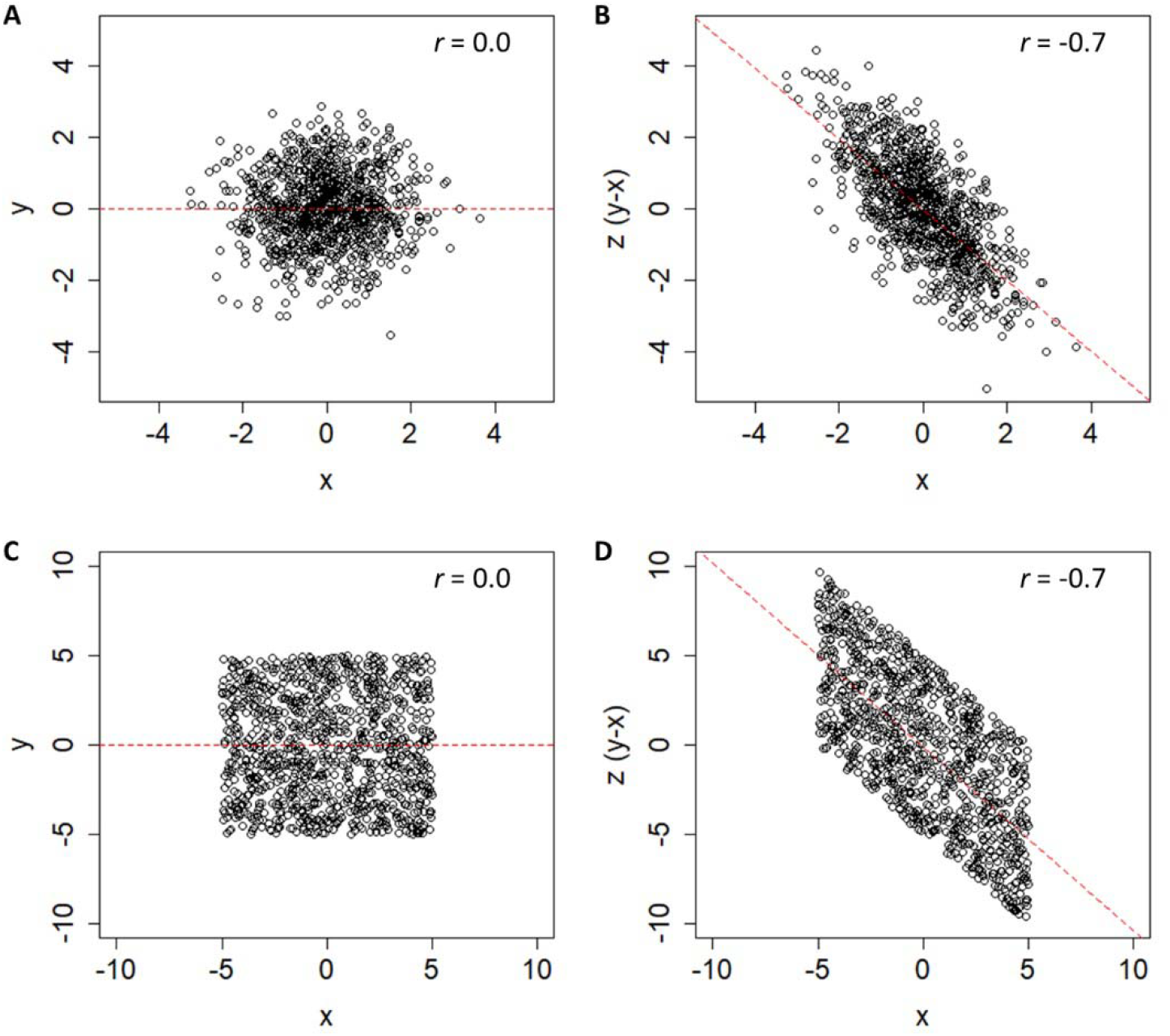
N = 1000 simulated data-points showing two uncorrelated variables, X and Y, and third variable Z, computed from their difference. In panels A and B, these variables are based on two normally distributed, but otherwise unbounded distributions. In panels C and D, these variables are based on two uniform distributions with bounds of ±5. In both cases, an artifactual negative relationship exists between X and Z, because those values are mathematically coupled.

As our “Change Score” (*Z*) is just our “Final Score” (*F*) minus our “Initial Score” (-*X*), it is not surprising that and are negatively related. In fact, the only thing distorting their relationship is *Y*. As Hope et al.^8^ pointed out, this is why the relative variance in *X* and *Y* matters. If the variance in *Y* is vastly *smaller* than *X*, we are essentially regressing *-X* onto *X*. If the variance in *Y* is vastly *bigger* than *X*, we are essentially regressing *Y* onto *X*.

As such, it is generally bad practice to regress change scores onto baseline scores; doing so will lead to relationships that are artifacts due to mathematical coupling, rather than genuine relationships (for other medical examples see^24-26^). Past critiques of proportional recovery have focused on the coupling that arises when change scores of the same variable are regressed onto baseline. However, it is important to point out this coupling also arises if we regress change scores onto other values that are correlated with our baseline assessment. For instance, if we regressed change in the FMA onto baseline Action Research Arm Test (ARAT) scores and found a significant relationship, that relationship might still be due to the fact that *baseline* FMA and ARAT scores are related, not that *change* in FMA is truly related to one’s baseline ARAT. To use a different variable, the same problem arises for corticospinal tract (CST) integrity: CST integrity is related to baseline FMA, so some proportion of its relationship to change in FMA is also likely due to mathematical coupling.

Our comments thus far have focused on how mathematical coupling is a general concern anytime change scores are regressed onto baseline characteristics. Now, we want to focus specifically on proportional recovery studies, where change in the FMA is regressed on baseline FMA scores. As shown in Figure 2, we have adapted the data gathered by Hawe et al.^7^ These data reflect several different available studies on proportional recovery using the Fugl-Meyer Assessment. ^2,14,27-30^ Regressing the N = 373 change scores onto baseline levels of impairment shows an overall slope of 0.42 when the non-fitters (as identified in past studies) are included in the data. If these non-fitters are excluded, then the slope of the regression line is shifted upward, to 0.76 (as shown by a dashed black line in Figure 2). In either case, this slope is statistically different from zero, *p’s* < 0.001. However, due to mathematical coupling any relationship we find is either a statistical artifact or at least inflated by such an artifact. As such, we need to consider the adequacy of a traditional hypothesis test here.

**Figure 2.**
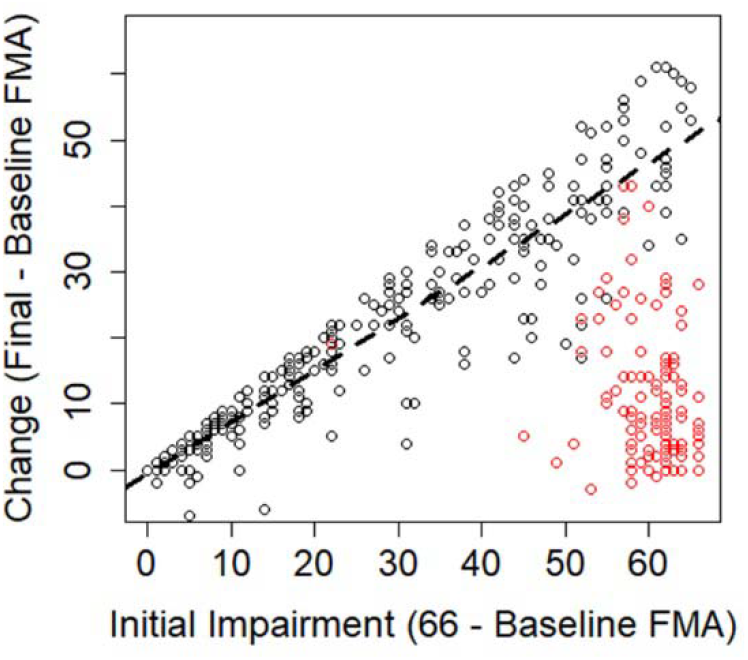
Data adapted from Hawe et al. (2019).^6^ Change scores and initial impairments extracted from empirical studies have been combined to create an “overall” sense of the relationship across studies. The dashed line denotes the ordinary least squared regression line for all fitters (black points). Data-points that were identified as non-fitters in the original studies are shown as red points.

## Consideration 2: Appropriate null hypotheses and alternative explanations

A common, null-hypothesis significance test for a regression slope assumes that the true value of the slope in the population is zero and that sampling variability is the only factor acting on the data. Other values can be chosen for the null hypothesis (e.g., H_0_: *β* = 0.5), but researchers often choose this “nil-hypothesis” significance test, where we explicitly assume the true effect is zero (i.e., the nil-hypothesis is a specific case of the null hypothesis where H_0_: *β* = 0, and for a two-sided test the alternative hypothesis would be H_a_: *β* ≠ 0). As we have shown in the previous section (and as past work has shown in detail^7,8^), it is not surprising to reject the nil hypothesis in this situation due to an artifact created by mathematical coupling. This artifact means that random sampling is not the only factor at work, nor should one expect a “true” relationship of zero, making the test of the nil-hypothesis H_0_: *β* = 0 uninformative.

It would, however, still be reasonable to ask if the observed relationship was greater than the mathematical artifact. This would require conducting a meaningful non-zero null-hypothesis test, and to do that the mathematical artifact needs to be estimated. We will turn our attention more to that issue in Consideration #3, but for now let us consider two situations in which the nil-hypothesis of *β* = 0 is rejected. In one case (Figure 3A), we have simulated *proportional* recovery that leads to an average level of recovery of about 50% (i.e., a regression slope of 0.50). In the other case (Figure 3B), we have simulated *random uniform* recovery which will always lead to an average level of recovery of about 50% (because approximately half of the data will be below/above the mean change at each level of impairment).

**Figure 3.**
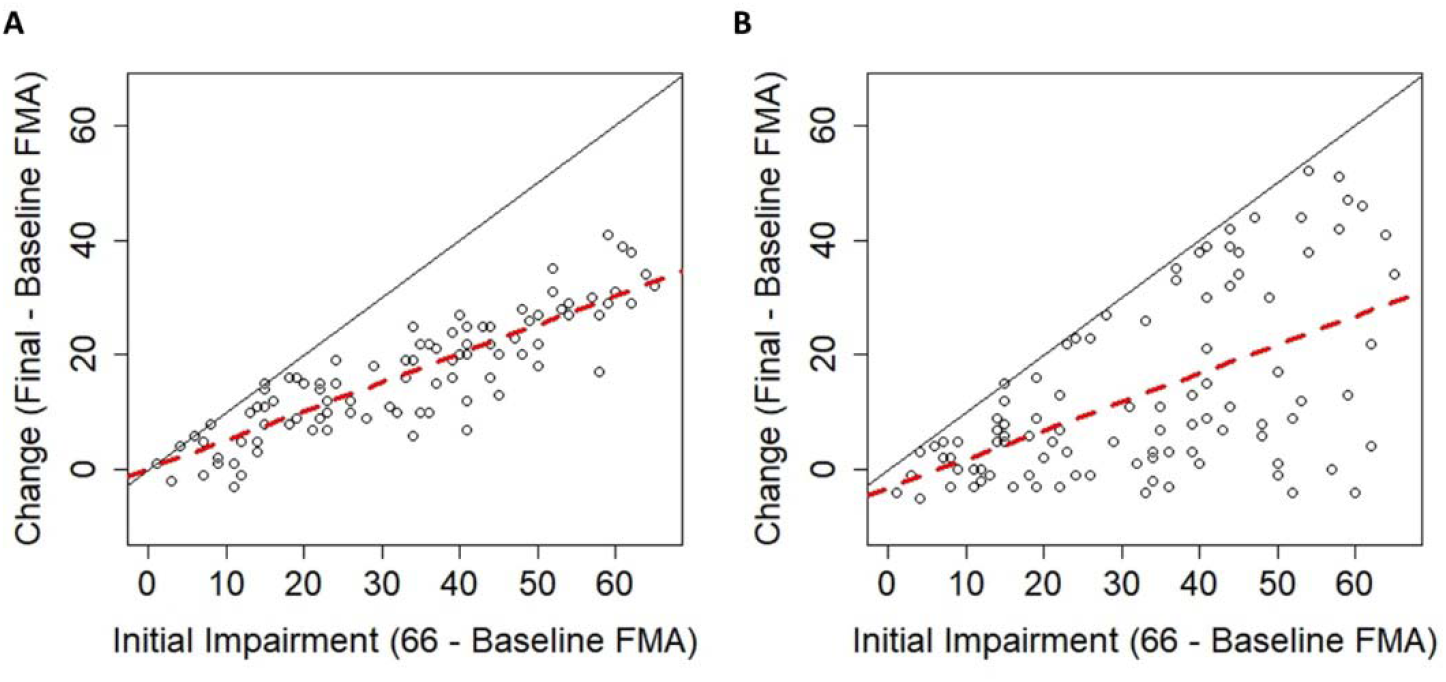
(A) Simulated data in which the change in Fugl-Meyer Assessment (FMA) is normally distributed around proportional recovery. (B) Simulated data in which the change in Fugl-Meyer Assessment (FMA) follows a uniform distribution. In both cases there is an upper bound on recovery due to the nature of the FMA (solid black lines), but even when change is random, there is a positive slope of ≈ 0.5 (dashed red lines).

For the proportional recovery example, data were normally distributed (σ = 6) around 50% of initial impairment, truncated by the ceiling of the FMA. Proportional recovery could be simulated with different parameters (e.g., other means/standard deviations), but this is meant to be only one example of proportional recovery. For the random uniform example, data are uniformly distributed between a slight negative change (−6 points) and the maximum points allowed on the FMA (maximum possible recovery). Again, the uniform distribution could have different parameters, but this is meant to be only one example of random uniform recovery to illustrate our point.

Random uniform recovery is a reasonable distribution in this situation, because it allows for the fact that different levels of recovery exist,^15,31,32^ but that the overall distribution of recovery covers the entire available space. Patterns like uniform recovery have been shown in animal data^33,34^ and human data using the FMA for the upper extremity^19^, the FMA for the lower extremity^3^, and the Arm Activity Measure^35^. It is important to note that modeling recovery as random uniform change does not mean that recovery is an inherently random process.

Consistent, distinct patterns of recovery almost certainly exist^19^ and are (at least) partially explained by physiological characteristics^2^. Random uniform recovery simply assumes that the full space of recovery is possible, and the distribution is uniform in the population (but naturally this will vary from sample to sample).

As we have shown, a proportional looking relationship already exists at the group-level whether recovery is proportional or uniform. In both cases in Figure 3 the regression slopes would suggest recovery is about 50% proportional and we would reject the nil hypothesis (H_0_: *β* = 0) in each case (proportional case: b=0.50, p<0.001; uniform case: b=0.49, p<0.001). Comparing the plots in Figure 3 helps to illustrate that rejecting this null hypothesis does not mean that a particular explanation for the effect is correct; it merely means that the data were unlikely to have arisen if the null were true. That is, we reject the null hypothesis, H_0_: *β* = 0, and we are inclined to accept the alternative, H_a_: *β* ≠ 0. However, rejecting the null does not imply that a particular theory/explanation/hypothesis is true. Clearly in the case of proportional recovery, it is neither the magnitude nor the statistical significance of the regression slope that make recovery proportional.

An important part of the proportionality argument is how individual changes are distributed around the group-level slope. If everyone is clustered around the group-level slope (Figure 3A), then the proportional argument seems very reasonable. Conversely, if everyone can be distributed across the entire space of recovery (Figure 3B), then we think this association is a statistical artifact (due to regressing bounded scales, change and initial impairment, onto each other). Contrasting the empirical data from Figure 2 against the simulated data in Figure 3, that conclusion crucially depends on whether or not “non-fitters” are included in the sample. In Consideration #3, we explore the validity of the fitters-classification by showing how hierarchical clustering procedures lead to the spurious identification of non-fitters even when recovery follows a random uniform distribution. Furthermore, in Supplemental Appendix II, we present analyses of the variability in the real data (N=373 shown in Figure 2) at different levels of initial impairment. In brief, those analyses show that the variance at most levels of initial impairment is not statistically different from random uniform recovery, but there are pockets of impairment levels where recovery shows less variance than expected.

Together, the simulation results raise questions about the validity of the fitters-classification and variance analyses suggest that **real** individual changes are not reliably different from uniform recovery at most levels of initial impairment (with notable exceptions in the middle of the distribution). Before we address those concerns, it is important to clarify what we mean by group-level statistics and individual-level data. As Kundert et al^4^ write in their abstract, “existing data are largely consistent with the [proportional recovery rule] at the population-level, […] recent reports of its demise are exaggerated, as they excessively focus on the less conclusive issue of individual subject-level predictions.”

When we are discussing individual-level data in this perspective, we mean how classification of individuals into fitters and non-fitters ultimately affects the group-/population-level statistics we observe when the data are aggregated. The decision about proportionality depends on how individuals spread around the group-level slope. Thus, the validity of excluding “non-fitters” is a critical issue. However, we do not mean making specific *predictions* about how an individual recovers. In the examples we have discussed so far, initial impairment is *associated* with*/explains* variance in change scores. We reserve the word “prediction *"* to refer specifically to the classification of individuals in independent samples (e.g., PREP^36^ or TWIST algorithms^37^). This sort of out-of-sample individual prediction was not the original purpose of the proportional recovery rule, nor is it part of our critique.

## Consideration 3: Measurement issues can create the illusion of proportionality

The fact that proportional recovery is apparent across many different scales of measurement has been argued as evidence for proportional recovery being a neurobiological phenomenon.^4^ First shown in the Fugl-Meyer Assessment (FMA^1^), proportional recovery has since been shown in the FIM^7^, the Western Aphasia Battery^37^ and the Letter Cancellation Test^38^, among other inventories. However, all these inventories possess lower and upper bounds. Although the individual minima and maxima are all different, the presence of these boundaries creates a real problem for interpreting the relationship between baseline scores and change scores. The code provided in the Supplemental Appendix I can be revised to demonstrate this point, but one can also consider Figure 3B in a thought experiment. Regardless of what the individual minima and maxima of these different scales are, random uniform recovery will always lead to the bottom triangle of the possible space being covered.

As shown in Figure 2, however, we can see that the story is more complicated than that because the distribution of initial impairments is not uniform. There are higher densities of very low and very high impairments on the Fugl-Meyer Assessment. Therefore, to make our simulations^40^ more realistic, we bootstrapped (i.e., repeatedly sampled) the initial impairment data from Hawe et al.^7^ (shown in Figure 2) to get a new “population” of 10,000 initial impairments with a similar distribution of initial impairments, but uniformly distributed change scores.

Using this simulated population, we can repeatedly draw samples to see how the data look from one sample to the next. One such sample of n=30 participants is shown in Figure 4A. The regression slope in this sample happens to be β=0.62, as shown by the dashed red line. Taking repeated samples, however, leads to a distribution of sample slopes as shown in Figure 4B. The dashed blue line shows the distribution of sample slopes from k=10,000 independent samples. As discussed above, these slopes vary around a population-level slope of β=0.50 purely due to the mathematical artifact of regressing change scores onto initial impairments. Thus, to decide if our single-sample slope of 0.62 is interesting, we need to take this mathematical artifact into account. As shown in Figure 4C, the slope of 0.62 is statistically surprising under the nil-hypothesis of no effect (p < 0.001; the dashed black line centered on 0), but is no longer statistically surprising when the null hypothesis takes the mathematical artifact from random uniform recovery into account (p = 0.337; the dashed blue line centered on 0.5). Large positive associations between change and initial impairment should be expected; not because of any inherent biological correspondence, but because of how we handled the data.

**Figure 4.**
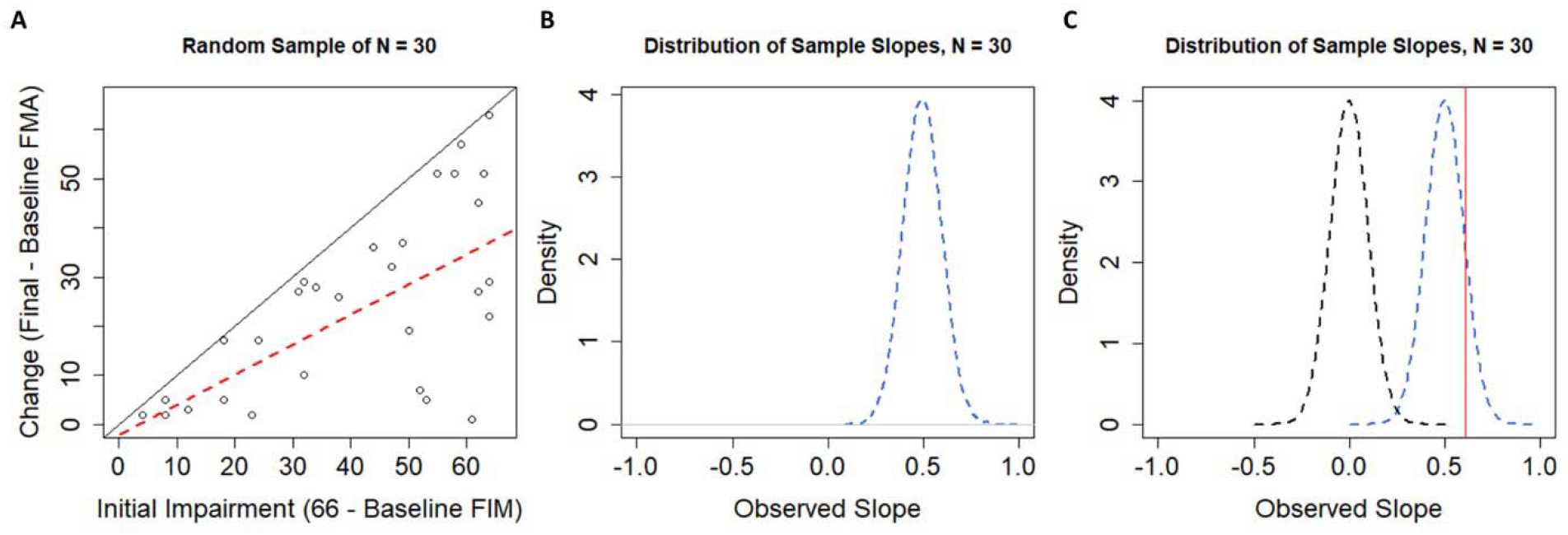
(A) A single random sample of N=30 FMA scores drawn from our population. The regression line (dashed red) has a slope of 0.62, which might suggest proportional recovery were it not drawn from a sample of randomly generated data. A diagonal black line with a slope of 1 is shown for reference. (B) The sampling distribution of slopes when our simulated population was sampled 10,000 times, with replacement, at sample sizes of N=30. (C) Contrasting the distribution of sample slopes under the null-hypothesis H_0_: β = 0 (dashed black line) and the distribution of sample slopes from our simulated population (centered on 0.5; dashed blue line). Note that now our observed slope of 0.62 (shown as the vertical red line) is no longer statistically significant when we use a null-hypothesis that takes the mathematical artifact into account (p<0.001, when H_0_: β = 0; versus p=0.337 when H_0_: β = 0.5).

At this point it is critical to consider the exclusion of “non-fitters” because that is how data are handled in proportional recovery studies. Methods that have been used to establish fitters from non-fitters can be legitimate methods whether they are data-driven methods (like hierarchical cluster analysis) or theory driven methods (such as moderator analyses using physiological data^2,15^). We are not debating the fundamental accuracy of these approaches, but their use in the classification of “fitters” and “non-fitters” in this context.

To address the validity of hierarchical clustering as a method for classifying individuals as fitters or non-fitters, we will use our simulated population of individuals with random uniform change scores. In the first run of our simulations, we took k=10,000 samples of N=30 individuals. In each sample, we used hierarchical cluster analysis^40,41^ to identify clusters of participants who could be classified as fitters and non-fitters. As shown in Figure 5A, the clustering algorithm still identifies clusters of participants as fitters (black dots) and non-fitters (red dots) when sampling from data with random uniform change scores. Obtaining clusters of fitters and non-fitters is obviously problematic in this case, because the underlying change is uniform. As such, we should be concerned that fitters and non-fitters may be (at least partially) an artifactual classification.

**Figure 5.**
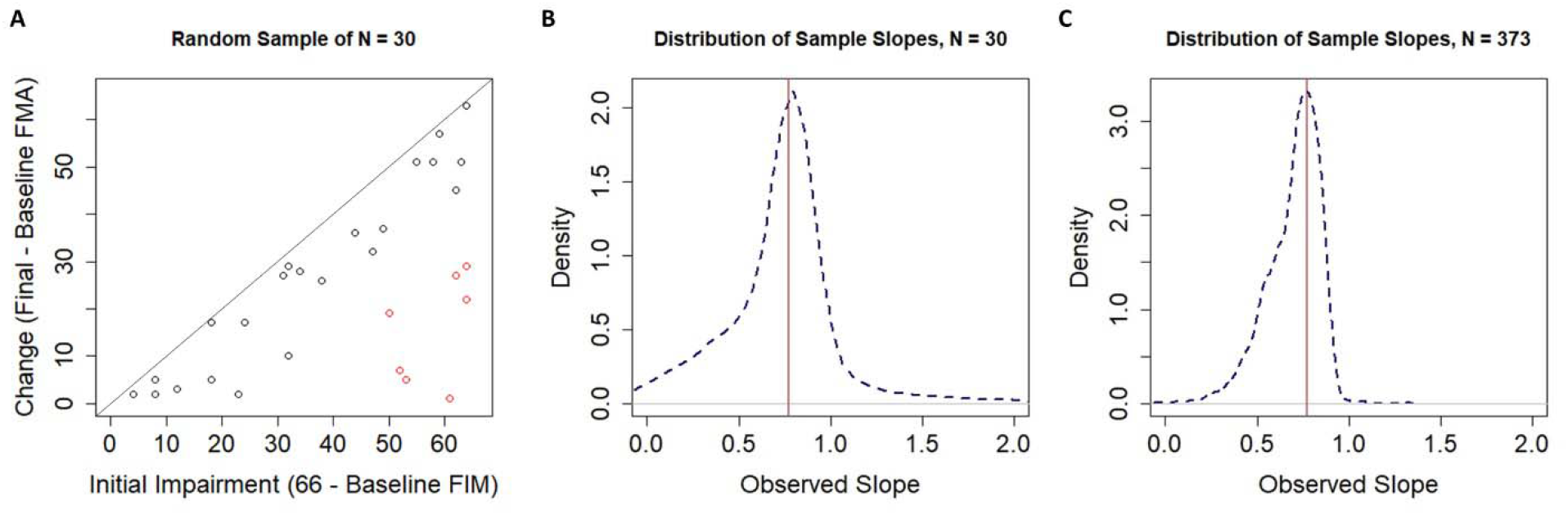
(A) A single random sample of N = 30 participants drawn from a population with random change scores. Note the clustering algorithm still classifies participants into what look like fitters and non-fitters even when there is no “rule” to which individuals can “fit”. (B) The distribution of sample slopes for fitters identified by our clustering procedure when the original sample size was N=30. (C) The distribution of sample slopes for fitters identified by our clustering procedure when the original sample size was N=373.

Current procedures described in studies of proportional recovery are not entirely clear on how their clusters were ascertained. That is, a cluster analysis can work using either a bottom-up agglomerative procedure or a top-down divisive procedure, but in general authors have an objective criterion for where they stop in determining their clusters. Although it has been stated that a criterion has been used^28^ it is not clear what the numeric value of this criterion is. ^2,29,38^ As such it is not clear by what criteria authors are making the decision to stop at two clusters in their analyses. We do know that authors are sometimes calculating Mahalanobis distances between points^42^ and that these distance values are being used as input into the agglomerative clustering algorithm advocated by Ward^41,43^. In the absence of a clear criterion by which we should stop clustering, we ran cluster analyses in our simulations that always stopped at two clusters. To determine which cluster was the “fitters”, we chose the cluster with a higher mean change score (consistent with the central argument of proportional recovery). Otherwise, our simulations used methods identical to published work for calculating distances and determining clusters.

As shown in Figure 5A, our clustering procedure leads to an identification of fitters and non-fitters to the proportional recovery rule consistent with past literature. Despite coming from a population of random uniform recovery, the clustering procedure spuriously identifies fitters (black dots) and non-fitters (red dots). This result is likely due to the asymmetry in the variable space. That is, when empirical and predicted change scores are fed into the algorithm, major deviations from the predicted change can only occur in the negative direction (lower right corner of Figure 5A), because individuals cannot improve beyond the maximum score of the scale (represented by the solid black line in Figure 5A). We show the effects of this procedure (sampling, clustering, and estimating slopes for the fitters cluster) when the total size is N=30 (to illustrate a relatively small, but common sample size) and when the total N=373, matching the total sample size for the pooled data.^7^

As shown in Figure 5B, when we simulated samples of size N=30, the distribution of sample slopes for the “fitters” had a mean of 0.770, a median of 0.781, and a negative skew. Thus, under this sampling distribution, we would not find it surprising to observe a large positive slope of 0.769 (as was observed in Hawe et al.^7^). Specifically, with a starting sample size of N=30, we’d expect a slope of ≥0.769 for the fitters about 54% of the time.

We reach a similar conclusion if we take the pooled real data for the fitters. The slope for the n=254 fitters out of those initial N=373 subjects was 0.769.^7^ As shown in Figure 5C, simulating random uniform recovery, hierarchical cluster analysis with two clusters, and that fitters would be the group with higher mean change, the mean of this sampling distribution was 0.778 and the median was 0.771. Based on this simulated distribution we would expect to get a slope of ≥0.769 for the fitters about 54% of the time.

Our simulated data (assuming random uniform recovery) led to a similar classification of “fitters” and “non-fitters” when fed into hierarchical clustering algorithms. (See Supplemental Appendix II for more details about these clusters.) This finding casts doubt on the validity of the fitters classification, because in the simulations there is no proportional recovery rule to which an individual can fit. This result suggests the fitters-classification is (at least in part) artifactual, likely due to the asymmetrical variable space. This spurious classification is a new finding and has important implications for how we should interpret other results.

First, data invoked as evidence for the proportional recovery rule are relatively weak. These patterns are quite consistent with what one might expect if recovery was uniformly and randomly distributed. Kundert et al.^4^ are quite correct when they wrote, “The fact that one can generate data that reproduces some findings of the PRR does not mean that the [proportional recovery rule] is invalid or that the observed data does not represent biologically meaningful associations.” However, if we had a null-hypothesis test p-value of p=0.54, we would not reject the null hypothesis. Proportional recovery is no different. We have shown that the group-level slope of b=0.769 carries a p=0.54 for fitters based on a sample of N=373 when we assume random uniform recovery. As such, we should not reject the hypothesis that recovery is uniformly distributed, nor should we accept the hypothesis that recovery is proportional at this time. Current data are consistent with both hypotheses, but the methods of measurement and analysis are rife with statistical limitations. Thus, we argue for abandoning current approaches to measuring proportional recovery, and we would generally caution against measuring recovery as the difference between two time points.

Second, studies showing that physiological characteristics are associated with the fitters/non-fitters classification should instead be reframed as physiological characteristics associated with important variation in recovery. For instance, individuals with poor cortico-spinal tract integrity are not “non-fitters”, they are just more likely to have significant impairment and poor recovery^2^. The integrity of specific brain regions clearly plays a role in the potential for recovery. There is, however, still substantial variability even among neuroanatomically similar individuals^32^. Recovery is a complex and multivariable problem and the field still has much work to do explaining individual differences in recovery trajectories.

## Discussion

The proportional recovery rule has been an influential finding in the field of neurorehabilitation. Recent debates about its accuracy and validity are also a very useful case-study, highlighting more general concerns for the study of recovery. Data argued to show proportional recovery in stroke rehabilitation have been found across a wide variety of assessments and replicated in many different samples. This pattern appeared so pervasive and the relationship so strong that it was compelling to think of this pattern as a neurological rule. As we have shown, however, current patterns claimed to be evidence for proportional recovery are generally consistent with uniform random recovery in these measures. This does not mean that proportional recovery has been disproven, but it does mean that we do not have the data to reliably say that recovery is proportional at this time.

Combining our simulations with analyses of the variance in the real data, our results cast doubt on the non-fitters classification and suggest variability in recovery is greater than what has been suggested by the proportional recovery rule. However, we also do not conclude that recovery is uniformly distributed across all levels of impairment (or for all measures). Indeed, as shown in Supplemental Appendix II, there are some levels of initial impairment where variance in change scores is lower than would be expected under random uniform recovery for the FMA. Thus, it is very likely there are non-uniformities, but it is not clear from the current data if this is due to non-linearities in the FMA or true systematic relationships between initial impairment and recovery. Ultimately, whether recovery is (partly) proportional or not, the methods commonly used to support proportional recovery are flawed and future arguments in favor of proportional recovery need to be based on different methods and data.

It is also important to remember that whether variation is uniform or not, the variation is not pure chance; it is uncertainty that needs to be explained. There is very compelling evidence that physiological variables explain individual differences in recovery^22^, but this is a very different question from claiming that recovery is proportional and that there are “non-fitters” to this general rule. As our simulations show, clusters of fitters and non-fitters emerge even when recovery in uniformly distributed. Thus, rather than concluding that individuals with lower cortico-spinal tract integrity are more likely to be “non-fitters”, we think a more appropriate conclusion is that individuals with lower cortico-spinal tract integrity are likely to be severely impaired and to show minimal recovery. There are reliable individual differences in recovery and people with more similar neuroanatomy following stroke are likely to show more similar patterns of recovery (although there is still variation in recovery trajectories for neuroanatomically similar individuals^32^).

### Limitations in Our Simulations

Our simulations used an empirical distribution of initial impairment values, but they assumed a uniform distribution of change scores for any given level of impairment. One could question the appropriateness of assuming random uniform change at-all, but especially in the context of the FMA upper extremity subscale. Visually, there appears to be an “island” of severely impaired individuals in the empirical data shown in Figure 2. This island is (at least) partially created by nonlinearities in the FMA^6,7^. Specifically, mid-range scores are less likely to occur in the FMA upper extremity subscale. If these mid-level scores are less likely, it makes sense that moderately impaired individuals will progress out of this range, but severely impaired individuals will be more likely to either surpass it or struggle to get into it (creating the island of non-fitters). Hawe et al.^7^ took this *bimodal* distribution of change scores into account in their simulations, but in the current study we explicitly chose to model change *uniformly* for four reasons.

First, the artifacts that are generated by regressing bounded change scores onto baseline scores are not a product of this nonlinearity (as shown in our simulations). Nonlinearity is a special concern for certain scales, but the problems created by ceiling/floor effects are more general. Second, data from both animal and human studies suggest that more uniform patterns of recovery exist for a variety of scales^3,19,33,39^. As such, uniform random recovery is a valid null hypothesis against which to test. Third, assuming uniform random recovery illustrates how using hierarchical cluster analysis with Mahalanobis distances can break down in this situation. When pairwise distances are calculated based on predicted change and actual change, these cluster analyses will spuriously identify fitters and non-fitters.^i^ Fourth, as shown in Supplemental Appendix II, this “island” is much less pronounced when recovery is normalized to the level of initial impairment and the observed variances in the real data are not statistically different from random uniform recovery, at most levels of initial impairment.

In our simulations, we did sometimes identify other types of clusters, especially at small sample sizes. We explored different methods for excluding these poorly ascertained clusters to see how it would affect the distribution of sample slopes for the fitters (e.g., rejecting samples where a cluster was less than 5% of the total sample size; rejecting samples where clusters were separated based purely on initial impairment). These steps affected the tails of the sampling distribution, but in all cases the distribution was centered near 0.7. Thus, although other processing decisions could have been made, of all of the processing decisions we explored, obtaining a slope of 0.7 for the fitters’ cluster was quite likely even when recovery was uniform and random.

### Beyond Proportional Recovery

The debate around proportional recovery also highlights questions about design, measurement, and statistical analysis that are broadly important to clinicians/researchers:

- **First, when designing a study, it is important to decide if our focus is on endpoints or trajectories**. If our focus is on endpoints, then endpoints should be our outcome and we should control for baseline measures as covariates. Conceptually this is more like Figure 6A, where we show the cumulative data from Hawe et al.^7^ Rather than plotting change as a function of initial impairment, we are now showing final FMA upper extremity scores as a function of baseline scores. If our focus is on trajectories, however, then we need to model change over time, more like Figure 6B. As we have shown, however, the distinction between fitters and non-fitters is, at least in part, an artificial classification and there could be many more groups that make up the possible recovery space, as shown conceptually in Figure 6C. To reliably model these longitudinal trajectories, however (e.g., in latent growth curve models, multi-level models), we need more than two data points. An advantage of these models is that they can avoid issues of mathematical coupling because the model estimates a trajectory rather than regressing change scores onto baseline variables. This is very much the technique adopted by van der Vliet et al.^19^ who used longitudinal mixture models to identify five subgroups of participants with a mean of 6.1 measurements per person. The insights gleaned from their longitudinal study show the power of these approaches, and we very much recommend these types of models (or similar^18,44,45^) as the field moves forward.
- **Second, it is important to remember that the null-hypothesis significance test answers a single, very specific question**^46,47^: “assuming all our assumptions are correct (the true effect is some specific value [often zero], sampling variability is the only factor acting on our data, etc.), what is the probability of observing data this extreme or more extreme?” Rejecting the null hypothesis means that we accept the alternative hypothesis in a statistical sense (i.e. we reject H_0_: *β* = 0; to favor H_a_: *β* ≠ 0), but it does not mean that a particular explanation of the effect is correct and we need to carefully consider how to choose between alternative explanations when the null is rejected.
- **Third, it is important to avoid subgrouping the data in an arbitrary manner**. Post-hoc segregation of change scores into fitters and non-fitters echoes similar pursuits like classifying “responders” and “non-responders” based on distributions of change scores^48^ and is similarly confounded^49^. Classifying individual responses to treatment is a very difficult proposition and it must be done carefully. Indeed, in most clinical trials, it might not even be possible^50^. That said, we also should not ignore reliable sub-groups/individual differences that are supported by outside information. Meaningful differences in recovery and initial impairment are related physiological variables^5,15^ and specific behavioral tests^6^. We stress again that we should not treat these relationships as evidence for non-fitters to an otherwise pervasive rule; we should think about recovery and impairment more continuously, with anatomy/physiology explaining important variation.
- **Fourth and finally, it is important to consider the properties (and limitations) of the scales that we are using to quantify recovery**. Many clinical scales have strong ceiling/floor effects, because healthy (“normal”) performance is the maximum/minimum against which performance is being measured. While this is a reasonable choice for scale design, we need to be very cautious about the effects of these boundaries^7,8^. Additionally, many clinical scales produce ordinal data, but we often treat these data as interval/ratio data (especially when aggregated). For instance, a +2-point change on the FMA could be due to a *single* two-point change in elbow extension, or to *two* one-point changes in shoulder flexion and pronation-supination. Thus, higher FMA scores generally mean less impairment, but two people with the same FMA score do not necessarily have same impairment, nor do differences in FMA scores always mean the same change in impairment. Treating ordinal data as interval data is not always a problem^51^ and there are times we might actually transform ordinal data into interval data^52^, but we always need to carefully consider the pros and cons of how we choose to measure “recovery”.

**Figure 6.**
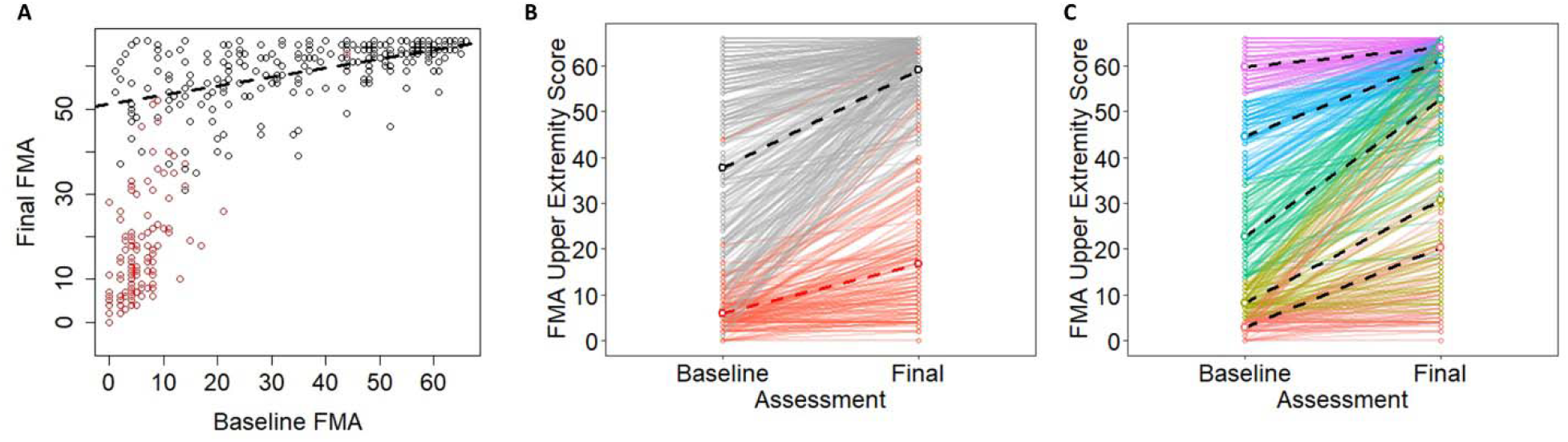
(A) Pooled empirical data from Hawe et al. shown as a function of baseline Fugl-Meyer Assessment scores. (B) The same data shown as trajectories for individual participants over time. Note that block dots correspond to “fitters” and red dots correspond to “non-fitters” in their original classifications. (C) A conceptual model in which the same data are color-coded based on quintiles of the baseline scores. Dashed lines show best fitting regression slopes within the various subgroups.

## Conclusions

Our goal in this point of view was to provide a “check-list” of conceptual and analytical issues in longitudinal measures of stroke recovery. We used proportional recovery as an illustrative example, but these issues of design, measurement, and analysis are broadly important for neurorehabilitation researchers. Using simulations, we showed: (1) how change scores can be problematic in this context; (2) how “nil” and non-zero null-hypothesis significance tests can be used and interpreted; and (3) how scale boundaries can create the illusion of proportionality (e.g., floor/ceiling effects), while other analytical procedures (e.g., spurious identification of non-fitters/non-responders) can augment this problem. Moving forward, understanding of the recovery process will be enhanced by embracing alternative designs (e.g., with more data collections at critical time-points), using different methods of analysis (e.g., that model true longitudinal trajectories), and exploring new outcome measures (e.g., that avoid the ceiling effects or ordinal, criterion-based scales).

## Data Availability

Currently, all empirical data underlying the simulations are available from the corresponding author upon reasonable request. All code for simulations are included in the supplemental appendix.

## Acknowledgments

The authors would like to thank Dr. Kristin Sainani, Dr. Thomas Hope, and three anonymous reviewers for their thoughtful comments on different drafts of this manuscript.

## Disclosure Statement

The authors received no funding specifically to pursue this work. SHS is the co-founder and Chief Scientific Officer of Kinarm that commercialize robotic technology for neurological assessment. All other authors have no conflicts of interest to declare.

## Notes

### Summary of Updates

Following reviewer comments, we have made numerous revisions throughout the text of the manuscript. Specifically, we have clarified the language around null and alternative hypotheses as distinct from scientific explanations. Additionally, we have added new analyses of the real data and present more details of our simulations in a new appendix: Supplemental Appendix II.

## References

1. Prabhakaran S, Zarahn E, Riley C, Speizer A, Chong JY, Lazar RM, Marshall RS, Krakauer JW. Inter-individual variability in the capacity for motor recovery after ischemic stroke. Neurorehabilitation & Neural Repair. 2008;22(1):64–71.

2. Byblow WD, Stinear CM, Barber PA, Petoe MA, Ackerley SJ. Proportional recovery after stroke depends on corticomotor integrity. Annals of Neurology. 2015;78(6):848–59.

3. Veerbeek JM, Winters C, van Wegen EE, Kwakkel G. Is the proportional recovery rule applicable to the lower limb after a first-ever ischemic stroke? PloS One. 2018;13(1):e0189279.

4. Kundert R, Goldsmith J, Veerbeek JM, Krakauer JW, Luft AR. What the Proportional Recovery Rule Is (and Is Not): Methodological and Statistical Considerations. Neurorehabilitation & Neural Repair. 2019;15:1545968319872996.

5. Stinear CM, Byblow WD, Ackerley SJ, Smith MC, Borges VM, Barber PA. Proportional motor recovery after stroke: Implications for trial design. Stroke. 2017;48 (3):795–8.

6. Senesh MR, Reinkensmeyer DJ. Breaking Proportional Recovery After Stroke. Neurorehabilitation & Neural Repair. 2019 Aug 16:1545968319868718.

7. Hawe RL, Scott SH, Dukelow SP. Taking Proportional Out of Stroke Recovery. Stroke. 2019;50(5):204–211.

8. Hope TM, Friston K, Price CJ, Leff AP, Rotshtein P, Bowman H. Recovery after stroke: not so proportional after all? Brain. 2019;141:15–22.

9. Cronbach, L. J., & Furby, L. (1970). How we should measure “change”: Or should we? Psychological Bulletin, 74(1), 68.

10. Linn, R. L., & Slinde, J. A. (1977). The determination of the significance of change between pre-and posttesting periods. Review of Educational Research, 47(1), 121–150.

11. Oldham PD. A note on the analysis of repeated measurements of the same subjects. J Chronic Dis. 1962; 15:969–977.

12. Gill JS, Zezulka AV, Beevers DG, Davies P. Relation between initial blood pressure and its fall with treatment. Lancet. 1985;1:567–569.

13. Tu YK, Gilthorpe MS. Revisiting the relation between change and initial value: a review and evaluation. Stat Med. 2007; 26:443–457. doi: 10.1002/sim.2538

14. Feng W, Wang J, Chhatbar PY, Doughty C, Landsittel D, Lioutas VA, Kautz SA, Schlaug G. Corticospinal tract lesion load: an imaging biomarker for stroke motor outcomes. Annals of Neurology. 2015;78(6):860–870.

15. Smith MC, Byblow WD, Barber PA, Stinear CM. Proportional recovery from lower limb motor impairment after stroke. Stroke. 2017;48(5):1400–1403.

16. Long JD. Longitudinal data analysis for the behavioral sciences using R. Sage; 2012.

17. Singer JD, Willett JB. Applied longitudinal data analysis: Modeling change and event occurrence. Oxford University Press; 2003.

18. Lohse K, Bland MD, Lang CE. Quantifying change during outpatient stroke rehabilitation: a retrospective regression analysis. Archives of physical medicine and rehabilitation. 2016;97(9):1423–30.

19. van der Vliet R, Selles RW, Andrinopoulou ER, Nijland R, Ribbers GM, Frens MA, Meskers C, Kwakkel G. Predicting upper limb motor impairment recovery after stroke: a mixture model. Annals of Neurology. 2020;87:383–393.

20. Burke QE, Dodakian L, See J, McKenzie A, Le V, Wojnowicz M, Shahbaba B, Cramer SC. Neural function, injury, and stroke subtype predict treatment gains after stroke. Annals of Neurology. 2015;77(1):132–45.

21. Stinear CM, Ward NS. How useful is imaging in predicting outcomes in stroke rehabilitation? International Journal of Stroke. 2013 Jan;8(1):33–7.

22. Stinear, CM, Smith, MC, Byblow, WD (2019). Prediction tools for stroke rehabilitation. Stroke. 2019;50:3314–3322.

23. Vickers AJ, Altman DG. Analysing controlled trials with baseline and follow up measurements. British Medical Journal. 2001;323(7321):1123–4.

24. Phang, P. T., Cunningham, K. F., Ronco, J. J., Wiggs, B. R., & Russell, J. A. (1994). Mathematical coupling explains dependence of oxygen consumption on oxygen delivery in ARDS. American Journal of Respiratory and Critical Care Medicine, 150(2), 318–323.

25. Tu, Y. K., Maddick, I. H., Griffiths, G. S., & Gilthorpe, M. S. (2004). Mathematical coupling can undermine the statistical assessment of clinical research: illustration from the treatment of guided tissue regeneration. Journal of Dentistry, 32(2), 133–142.

26. Browne JP, van der Meulen JH, Lewsey JD, Lamping DL, Black N. Mathematical coupling may account for the association between baseline severity and minimally important difference values. Journal of clinical epidemiology. 2010;63(8):865–74.

27. Buch ER, Rizk S, Nicolo P, Cohen LG, Schnider A, Guggisberg AG. Predicting motor improvement after stroke with clinical assessment and diffusion tensor imaging. Neurology. 2016;86(20):1924–1925.

28. Guggisberg AG, Nicolo P, Cohen LG, Schnider A, Buch ER. Longitudinal structural and functional differences between proportional and poor motor recovery after stroke. Neurorehabilitation & Neural Repair. 2017;31(12):1029–41.

29. Winters C, van Wegen EE, Daffertshofer A, Kwakkel G. Generalizability of the proportional recovery model for the upper extremity after an ischemic stroke. Neurorehabilitation & Neural Repair. 2015;29(7):614–22.

30. Zarahn E, Alon L, Ryan SL, Lazar RM, Vry MS, Weiller C, Marshall RS, Krakauer JW. Prediction of motor recovery using initial impairment and fMRI 48 h poststroke. Cerebral Cortex. 2011;21(12):2712–21.

31. Lin DJ, Cloutier AM, Erler KS, … Cramer SC. Corticospinal tract injury estimated from acute stroke imaging predicts upper extremity motor recovery after stroke. Stroke. 2019;50:00–00. DOI: 10.1161/STR0KEAHA.119.025898.

32. Findlater SE, Hawe RL, Mazerolle EL, … Dukelow SP. Comparing CST lesion metrics as biomarkers for recovery of motor and proprioceptive impairments after stroke. Neurorehabilitation & Neural Repair. 2019;1-14. DOI: 10.1177/1545968319868714

33. Jeffers MS, Karthikeyan S, Corbett D. Does stroke rehabilitation really matter? Part A: proportional stroke recovery in the rat. Neurorehabilitation & Neural Repair. 2018;32(1):3–6.

34. Jeffers MS, Karthikeyan S, Gomez-Smith M, Gasinzigwa S, Achenbach J, Feiten A, Corbett D. Does stroke rehabilitation really matter? Part B: An algorithm for prescribing an effective intensity of rehabilitation. Neurorehabilitation & Neural Repair. 2018;32(1):73–83.

35. Ward NS, Brander F, Kelly K. Intensive upper limb neurorehabilitation in chronic stroke: Outcomes from the Queen Square programme. J Neurol Neurosurg Psychiatry. 2019;90:498–506.

36. Stinear CM, Byblow WD, Ackerley SJ, Smith MC, Borges VM, Barber PA. PREP2: A biomarker based algorithm for predicting upper limb function after stroke. Annals of Clinical and Translational Neurology. 2017 Nov;4(11):811–20.

37. Smith MC, Barber PA, Stinear CM. The TWIST algorithm predicts time to walking independently after stroke. Neurorehabilitation & Neural Repair. 2017 Oct;31(10-11):955–64.

38. Lazar RM, Minzer B, Antoniello D, Festa JR, Krakauer JW, Marshall RS. Improvement in aphasia scores after stroke is well predicted by initial severity. Stroke. 2010;41(7):1485–8.

39. Winters C, Van Wegen EE, Daffertshofer A, Kwakkel G. Generalizability of the maximum proportional recovery rule to visuospatial neglect early poststroke. Neurorehabilitation & Neural Repair. 2017;31(4):334–42.

40. R Core Team (2019). R: A language and environment for statistical computing. R Foundation for Statistical Computing, Vienna, Austria. https://www.R-project.org/

41. Murtagh F, Legendre P. Ward’s hierarchical agglomerative clustering method: which algorithms implement Ward’s criterion? Journal of Classification. 2014;31 (3):274–95.

42. McLachlan GJ. Mahalanobis distance. Resonance. 1999;4(06).

43. Ward Jr JH. Hierarchical grouping to optimize an objective function. Journal of the American Statistical Association. 1963;58(301):236–44.

44. Kozlowski AJ, Heinemann AW. Using individual growth curve models to predict recovery and activities of daily living after spinal cord injury: an SCIRehab project study. Archives of Physical Medicine and Rehabilitation. 2013 Apr 1;94(4):S154–64.

45. Garcia TP, Marder K. Statistical approaches to longitudinal data analysis in neurodegenerative diseases: huntington’s disease as a model. Current Neurology and Neuroscience Reports. 2017 Feb 1;17(2):14.

46. Greenland, S., Senn, S. J., Rothman, K. J., Carlin, J. B., Poole, C., Goodman, S. N., & Altman, D. G. (2016). Statistical tests, P values, confidence intervals, and power: a guide to misinterpretations. European journal of epidemiology, 31(4), 337–350.

47. Sainani, K. L. (2009). Putting P values in perspective. PM&R, 1(9), 873–877.

48. Dankel SJ, Loenneke JP. A method to stop analyzing random error and start analyzing differential responders to exercise. Sports Med. 2019. https://doi.org/10.1007/s40279-019-01147-0.

49. Tenan, Vigotsky, & Caldwell (2019). Comment on: “A Method to Stop Analyzing Random Error and Start Analyzing Differential Responders to Exercise” Sports Medicine. 2019;50:431–434.

50. Senn S. Individual response to treatment: is it a valid assumption? BMJ. 2004;329:966–968.

51. Knapp, T. R. Treating ordinal scales as interval scales: an attempt to resolve the controversy. Nursing Research. 1990;39(2):121–123.

52. Tennant, A., Geddes, J. M., & Chamberlain, M. A. The Barthel Index: an ordinal score or interval level measure?. Clinical Rehabilitation, 1996;10(4):301&308.

